# An observational study of survival outcomes of people referred for ‘fast-track’ end-of-life care funding in a District General Hospital; too little too late?

**DOI:** 10.1101/2022.12.23.22283895

**Authors:** Jo Morrison, Cherry Chowdhary, Ryan Beazley, James Richards, Charlie Davis

## Abstract

**Background:** End-of-life care frequently requires support for people to die where they feel safe and well-cared for. End-of-life care may require funding to support dying outside of hospital. In England, funding is procured through Continuing Healthcare Fast-Track funding, requiring assessment to determine eligibility. Anecdotal evidence suggested that Fast-Track funding applications were deferred where clinicians thought this inappropriate due to limited life-expectancy.

**Aim:** To evaluate overall survival after Fast-Track funding application.

**Design:** Prospective evaluation of Fast-Track funding application outcomes and survival.

**Setting/participants:** All people in 2021 who had a Fast-Track funding application from a medium-sized district general hospital in Southwest England.

**Results:** 439 people were referred for Fast-Track funding with a median age of 80 years (range 31-100 years). 413/439 (94.7%) died during follow up, with a median survival of 15 days (range 0 to 436 days). Median survival for people with Fast-Track funding approved or deferred was 18 day and 25 days, respectively (P= 0.0056). 103 people (29%) died before discharge (median survival 4 days) and only 8.2% were still alive 90 days after referral for Fast-Track funding.

**Conclusions:** Fast-Track funding applications were deferred for those with very limited life-expectancy, with minimal clinical difference in survival (7 days) compared to those who had applications approved. This is likely to delay discharge to preferred place of death and reduce quality of end-of-life care. A blanket acceptance of Fast-Track funding applications, with review for those still alive after 60 days, may improve end-of-life care and be more efficient for the healthcare system.

**WHAT IS ALREADY KNOWN ABOUT THE TOPIC?:**

- People approaching the end-of-life may have rapidly deteriorating or fluctuating care needs, requiring a responsive care package to optimise care.
- Time to put in place care packages, to enable people to die in their preferred place, may be limited and so systems to facilitate care should be provided at speed.
- Continuing Healthcare Fast-Track (CHCFT) funding was designed to deliver person-centred care for people with ‘rapidly deteriorating condition, and where that condition may be entering a terminal phase’ without a specific measure of deterioration rate or prognostic expectation.

**WHAT THIS PAPER ADDS:**

- When clinical teams refer for CHCFT they are highly likely to be identifying someone who is in the last few days to weeks of life.
- Referral deferment (rejection) may correlate with survival statistically, but this was not a clinically meaningful difference.
- Local CHCFT eligibility interpretation inappropriately excluded people who need funding to be looked after in their preferred place of care in their last days of life.

**IMPLICATIONS FOR PRACTICE, THEORY OR POLICY:**

- The current application process for funding may prevent rapid discharge to preferred place of care for those with only a few days to live.
- A blanket policy of acceptance of care needs, with review at 60 or 90 days if still required, may improve quality of end-of-life care for people and their families, and may have cost savings to the health and social care system as a whole.

## BACKGROUND

End-of-life care is important for the wellbeing of people who are dying and the longer-term wellbeing of their surviving family and friends. This is a situation where there is only once chance to get it right and failures can lead to poor quality end-of-life care and contribute to abnormal grief responses in those bereaved, leading to ongoing costs to the bereaved and the wider economy.

The need to promote high-quality care for all adults at the end of life was highlighted in the UK Department of Health’s 2008 End of Life Care Strategy.^1^ However, there are significant variabilities in the quality of care and place of death nationally.^2^ Although many people with advanced illness would choose to die at home, in reality the majority in the UK spend their last moments in hospital, although the proportion of deaths at home increased between 2004-2010. ^3-7^

In England, the Continuing Healthcare Fast-Track (CHCFT) pathway was designed to enable urgent provision to aid people dying, in order to assist them in receiving appropriate support, either in their own home or in a care setting.^8^ That the individual has a ‘rapidly deteriorating condition and the condition may be entering terminal phase, is in itself sufficient to establish eligibility.’ The National Framework for National Health Service (NHS) Continuing Healthcare and NHS-funded Nursing Care report found a total of 53,745 people eligible for NHS CHC funding on the last day of Q4 2021-22. Of these, and 20,008 were eligible for fast-track care.^9^ Appropriately timed CHCFT decisions, together with proactive advance care planning and treatment escalation plans helps individuals to stay in the community, if that is their preference, avoiding or reducing the length of acute hospital admissions during the terminal phase of an illness, unless admission is required for management of uncontrolled symptoms.^10^

An initial step for a fast-track referral for end-of-life care is prognosis prediction, often significantly over-estimated;^11^ in one study survival from clinician estimates was only 25 days, whereas clinicians estimated 75 day median survival, and disclosed 90 day prognosis to their patients.^12^ Data from Germany found that median overall survival after discharge to the community for end-of-life care from specialised inpatient palliative care or other inpatient care settings was 24.0 days (range 1 to 488 days) for a cohort of 245 people.^13^ Most people were discharged to their own home (60.8 %), 20.0% to hospices (20.0 %) and 11.0% to nursing homes (11.0 %) and more than half remained in their preferred discharge setting (55.9%). However, the other 44.1 % of people had an average of 3.1 (± 4.1) changes of care setting; from home to hospital (32.4 %) and from hospital back to private home (24.4 %). This demonstrates often rapidly changing care needs for this cohort of people, who therefore need correspondingly rapidly responsive delivery of care.

CHCFT criteria specifically include those with minimal symptoms in whom a ‘rapid deterioration is to be expected in the near future’. CHCFT guidelines suggest review of care needs and eligibility at three months and again at least 12-monthly. Unfortunately, CHCFT care often appears to be restricted to those with less than two to three months to live. This may be due to misinterpretation by healthcare teams and those funding care. Lack of earlier CHCFT care packages may result in delays in acute hospital discharge, or inappropriate hospital admissions where care needs deteriorate rapidly, affecting the quality of end-of-life care for individuals and their families and friends, which may have long term impact on responses to bereavement.

Our hospital Trust encompasses acute and community care settings, including community hospitals and district nursing teams. At our Trust end-of-life steering group meeting, concerns were raised about individuals whose discharge to home was either delayed until last few days or hours of life, or not possible because they became so unwell that death was imminent. A basic tenant of quality improvement methodology, often misattributed to W. Edwards Deming, is ‘in God we trust, others must provide data’.^14^ We therefore sought to determine whether this anecdotal evidence represented reality, or whether this was special cause variation, in order to inform county-wide end-of-life care provision.

## METHODS

### Description of the data and the population

This audit was based in a district general hospital serving a local population of 350 000 and specialist services to a wider population of 800 000. The population served is significantly older than the UK average with double the UK average of over 65s and over 80s.^15^ Data for all patients referred to the CHCFT discharge team were collected prospectively onto a Microsoft Excel database,^16^ including date of referral, date of discharge and preference for place of delivery of end-of-life care, as part of routine tracking and fail-safe of care pathways.

### Measures

Patients referred for CHCFT end-of-life care in 2021 were followed up via their hospital electronic health records (EPRO^17^) to look for information for date of death and/or evidence of clinical activity following the CHCFT referral submission.

### Analysis

Data were analysed with Microsoft Excel^16^ and Prism^18^ and Kaplan-Meir plots generated and groups compared with Log-rank (Mantel-Cox) and Gehan-Breslow-Wilcoxon tests, as appropriate. Data were tested for normality and non-parametric datasets were analysed using Kruskal-Wallis and Mann-Whitney tests. Performance outcomes data were analysed with P-charts using LifeQI.^19^

### Ethics

The Somerset NHS Trust research ethics committee deemed ethics approval was not required, after completion of the Health Research Authority decision tool (http://www.hra-decisiontools.org.uk/research/) as this was a service evaluation of outcomes of clinical care and part of a wider “Last 1000 days” quality improvement programme.

## RESULTS

During 2021 there were 439 separate referrals for fast-track funding for end-of-life care placement. Median age for the cohort was 80 years (range 31-100). There was no difference in age of those whose fast-track funding was approved, deferred of those who died on the ward when compared to the entire cohort (median age 82, 78, 79.5 years, respectively; P = 0.23) (**Figure 1**). However, direct comparison of those approved and deferred demonstrated those deferred were younger (P= 0.047).

**Figure 1.**
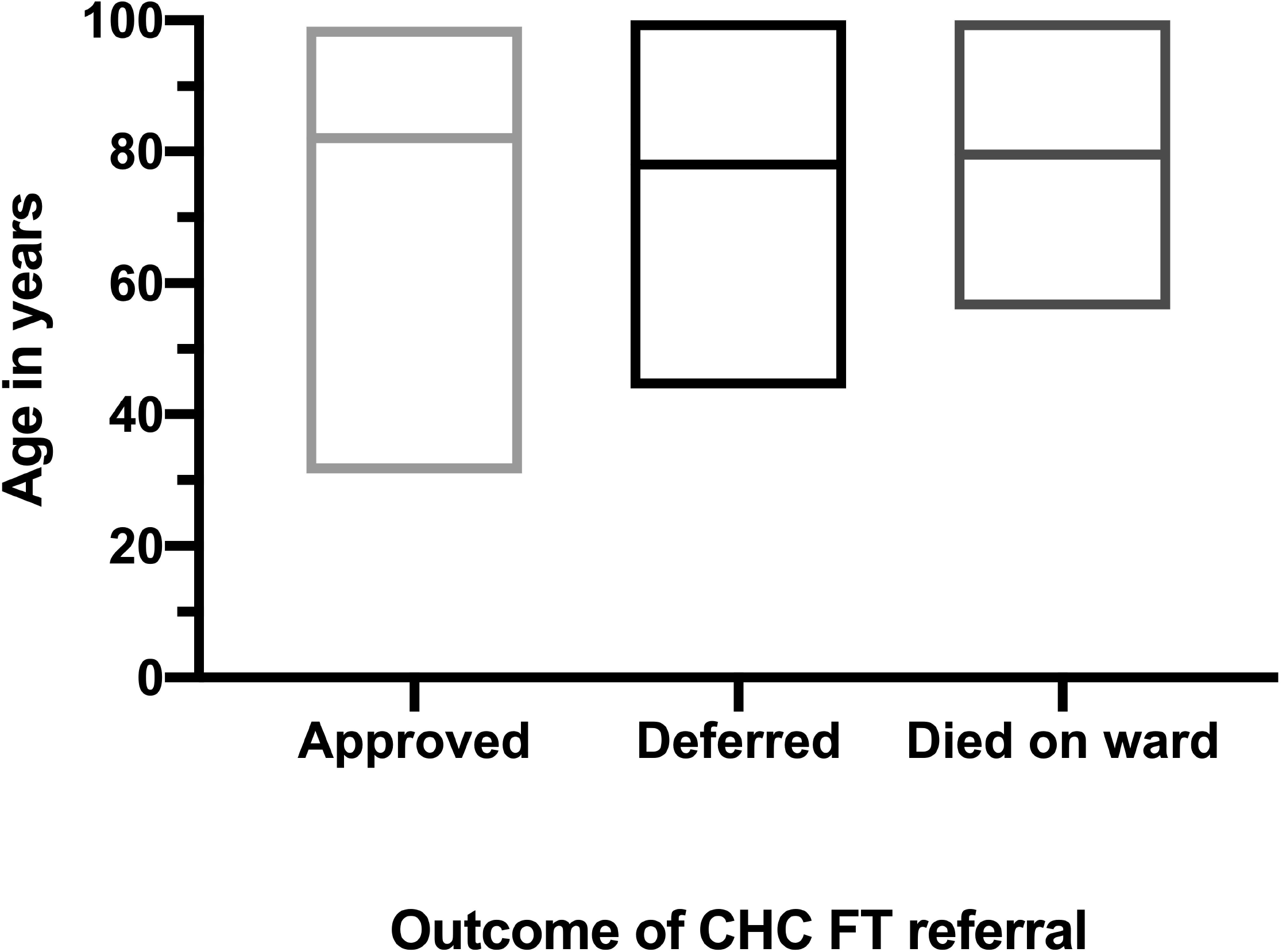
Age in years at referral for CHCFT funding. Bar at median age per group and box at minimum and maximum ages.

Two hundred and eight people (47.4%) were discharged with an approved funding application, 128 (29.2%) had funding deferred and 103 (23.4%) died whist still an inpatient. From the cohort, date of death was determined for 413 people at the time of follow up (6 to 18-month follow-up from referral depending on month of initial referral). The other 26 people were censored at the last known time they were alive, based on hospital records (subsequent admissions, attendance at outpatient appointments, blood test results in the community). One person was lost to follow up as they were discharged for end-of-life care to a nursing home out of area, so censored at the time of discharge.

Median survival for the entire cohort was 15 days (range 0 to 436 days) from the initial decision to refer from fast-track funding when it was recognised that the person was within the last weeks to months of life (**Figure 2**). There was a difference in median survival between those whose funding was approved and those whose funding was deferred (18 versus 25 days; P= 0.0056), but this was by only seven days. From the entire cohort, regardless of funding outcome, 56 of 439 people (14%) survived more than 60 days, and 33 people (8.2%) were still alive at 90 days from referral. Even for those who had funding deferred, as they were thought not to meet criteria of rapid decline and poor prognosis, only 26 of 128 people (23.4%) survived beyond 60 days. Nearly one third of the cohort (103 people; 29%) died on the ward due to rapid decline and/or personal/family choice, with a median survival four days. Between February to August there was a significant downward trend fast-track funding approval rates (**Figure 3**).

**Figure 2.**
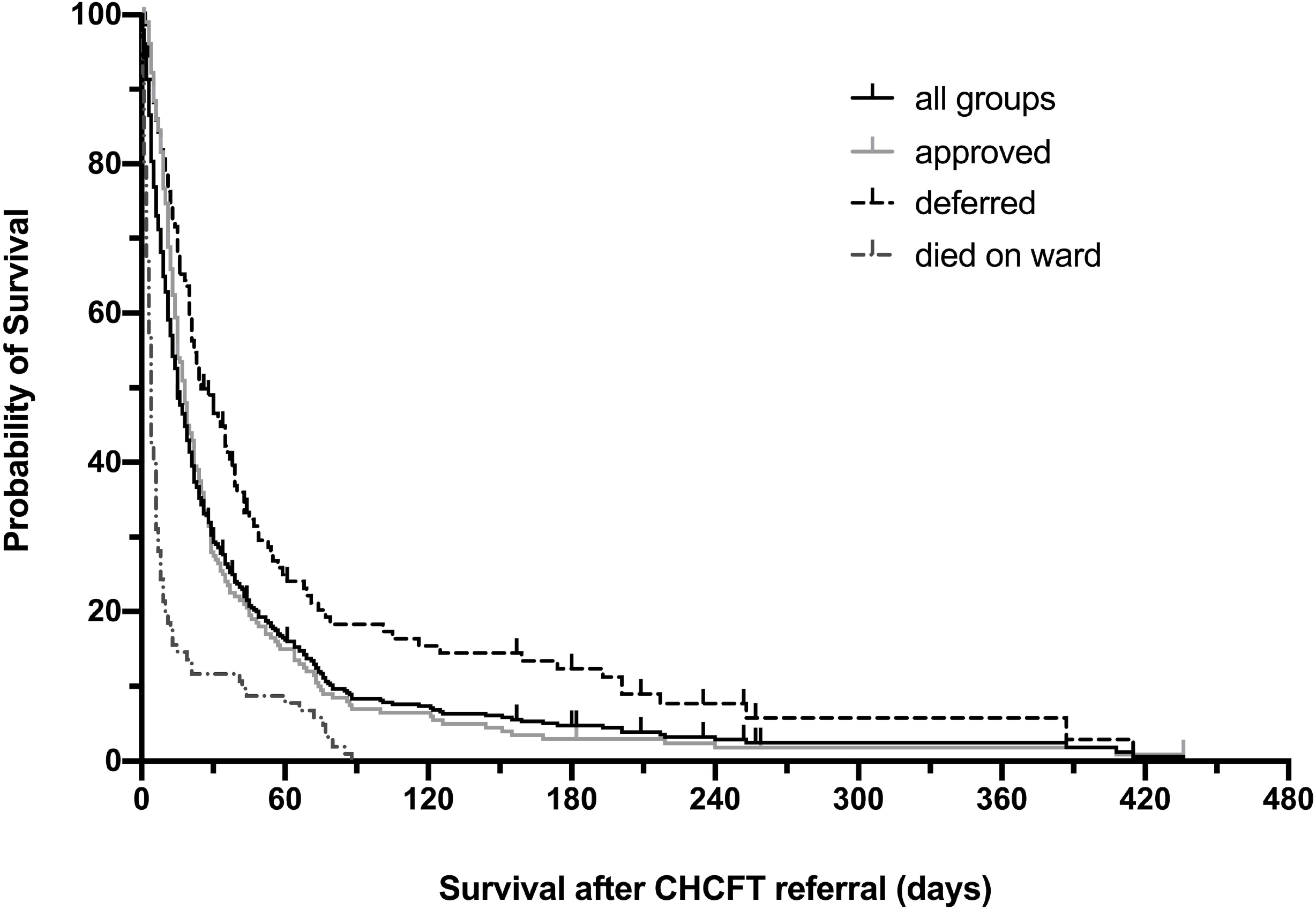
Kaplan-Meier plot of survival over time by outcome of CHCFT referral. Outcomes for entire cohort (black, solid line), approved (light grey, solid line), deferred (dark grey dashed line) and those that died on the ward (mid-grey, dash-dot line).

**Figure 3.**
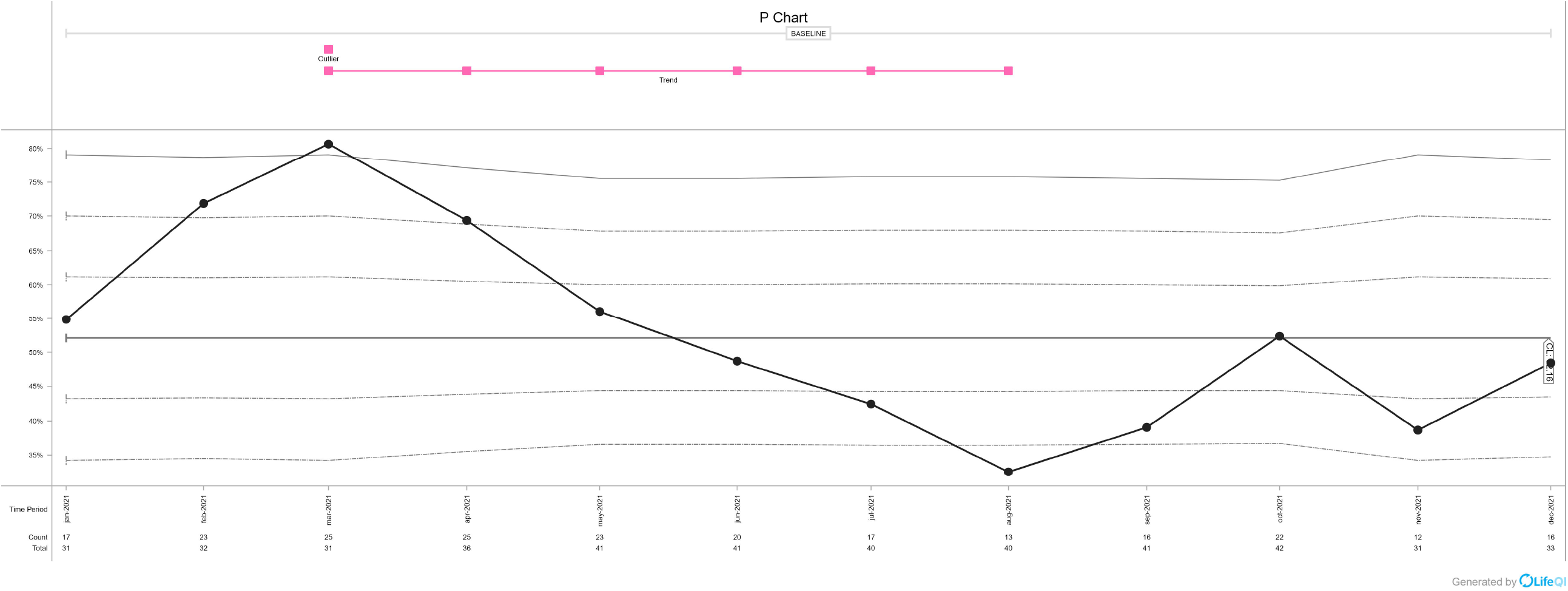
Statistical Process Control (SPC) chart (P Chart) of percentage of CHCFT referrals approved, by month of referral, over time. Median approval rate 52.16%. Significant downward trend in approval rates from March 2021 to August 2021 (following the local peak in Winter 2020-21 COVID-19 admissions).

## DISCUSSION

### Main findings/results of the study

People referred for end-or-life care packages during an inpatient admission to a district general hospital were identified in their last few days to weeks of life. Regardless of the decision on funding, few people were alive after 60 days and only 8.2% were alive after 90 days. Many patients were in their last few days to weeks of life and many did not survive long enough, or were not well enough, to be discharged to home, hospice or a nursing home. Over the audit period, the approval rate of funding declined. These analyses suggest that most people fulfilled CHCFT requirements at referral of anticipated poor prognosis and rapid decline.

### What this study adds

The study demonstrates that hospital staff identified people were approaching their last few days of life. However, despite this, placement funding was often deferred, as people were not thought to have had a ‘rapid deterioration’ in their condition, despite their condition warranting inpatient admission.

One suggestion from this work is that time and resources would be better spent rapidly delivering emergency end-of-life care for all thought to have a severely limited prognosis and to reassess funding decisions at 90 days, as per the guidelines, if the individual was still alive and likely to live longer than another few weeks at that point. This was especially relevant in 2021, as for much of that year there was very limited visiting in hospital due to the COVID-19 pandemic. It is interesting to note that approval rates declined as the winter peak of the COVID-19 pandemic waned.

The impression from review of the patient cohort, was that it appeared to be more challenging to determine prognosis in those with underlying frailty of old age, cardiac failure and COPD compared to those with advanced cancer. This hypothesis was not formally tested. Many people within our cohort, had at least one, if not several, admissions in the few months leading up to the admission which precipitated the fast-track funding referral. This may indicate that opportunities for advance care planning and treatment escalation plans were missed, with people referred to an acute hospital setting when alternative care may have been in their best interest, had advance care planning conversations been had at an earlier stage.

### Strengths and weaknesses/limitations of the study

A strength of this study is the prospective audit of people referred for fast-track funding and identification those who had referrals deferred as well as those approved. Another strength of the study is that we have a relatively defined and settled population, with the Trust spanning both acute and community hospitals in the area, and so only one person was lost to follow up.

However, a major limitation is that we were limited to including those who had referrals made in the acute patient setting. We do not have an electronic patient record that spans primary, community and acute care, and so we were not able to see if previous referrals had been made (and deferred) in the community prior to admission, nor ready access of clinicians to community advance care plans. This wider overview would better inform some of our inferences.

## CONCLUSION

CHCFT funding referrals in our acute hospital were made for people who were in the last few days to weeks of life. Nearly one third were so near to end-of-life that there was minimal opportunity to consider other settings for end-of-life care, which may have been their preference. Changes to how CHCFT referrals are processed, with an assumption that the clinicians have correctly identified that people have very little time remaining, might reduce the number of people who died in hospital. Furthermore, earlier identification of the terminal phase of illness, and better advance care planning, might prevent hospital admissions, allowing people to die with appropriate support at home or in nursing homes nearer to their families.

## Data Availability

Data, without patient identifiers, available on reasonable request from the authors.

## ACKNOWLEDGEMENTS

We thank:

The hospital fast-track discharge team led by Helen Greene, Tom Brown and Julie Maddock for their help with data collection, and members of the Hospital End of Life Steering group for raising concerns about CHCFT funding decisions, which lead to the idea for this study.

## AUTHORSHIP

JM and CD developed the original idea for the study. JM, JR, CC and RB performed further data collection. JM performed data analysis and JM, CD, JR, CC and RB contributed to writing and approved the final version of the paper.

## COMPETING INTERESTS

None

## FUNDING

None

## DATA REQUESTS

Data, without patient identifiers, available on reasonable request from the authors.

## Notes

### Competing Interest Statement

The authors have declared no competing interest.

### Funding Statement

This study did not receive any funding

### Author Declarations

Ethics committee/IRB of Somerset NHS Foundation Trust waived ethical approval for this work

